# Frailty is associated with low physical activity and poor sleep quality in patients undergoing myeloablative allogeneic hematopoietic cell transplantation: A Fitbit® Pilot Study

**DOI:** 10.1101/2024.03.27.24304863

**Authors:** Caryn R. Libbert, Fiona He, Najla El Jurdi, Helen Fagrelius, Mark Juckett, Joseph Maakaron, William Juckett, Nicholas Evanoff, Donald R. Dengel, Shernan G. Holtan

## Abstract

Allogeneic hematopoietic cell transplantation (alloHCT) is a vital therapy for various hematologic diseases, though it demands high physiological resilience. Frailty, a syndrome impacting the body’s ability to withstand stress, affects outcomes of alloHCT across all ages.This study examines the relationship between frailty and peri-transplant activity and sleep patterns using Fitbit® devices. In this pilot study, adults scheduled for their first myeloablative alloHCT at the University of Minnesota from June 2022 to January 2023 were included if they had a compatible device for the Fitbit® app. Participants were monitored for activity and sleep from admission to day +30 post-transplant. Frailty was assessed pre-transplant using Fried Phenotype criteria. Data were analyzed for activity and sleep patterns differences among not frail, pre-frail, and frail groups. Nine patients provided sufficient data for analysis, showing significant variances in activity levels and sleep patterns across frailty categories. Not frail patients exhibited significantly higher daily steps and active minutes than pre-frail and frail patients. Not frail patients also had the highest amount of restorative deep and rapid eye movement sleep. Due to Fitbit methodology and likely frequent interruptions, 28% of the days in the first month post-transplant had a recorded sleep time of 0 minutes. Although our sample size was small, our findings underscore the importance of frailty in influencing activity and sleep patterns among alloHCT recipients.

## INTRODUCTION

Allogeneic hematopoietic cell transplantation (alloHCT) is a critical treatment for various hematologic conditions, replacing diseased marrow with healthy stem cells from a donor. Despite its effectiveness, alloHCT demands significant physiological resilience from patients. Frailty, a syndrome characterized by a reduction in the body’s physiologic reserve—the ability to withstand stress and repair tissue damage—is not restricted to older individuals. It also affects younger patients undergoing alloHCT because of underlying disease or treatment-related toxicities, leading to an increased risk of mortality and post-transplant complications (1-3). Frailty presents in physical, psychological, and social dimensions, affecting patients’ ability to cope with and recover from alloHCT. Recognizing the importance of monitoring patient well-being during treatment, this study employs Fitbit® devices to measure activityx and sleep patterns—key health indicators during the peri-transplant period (4). While research has established the detrimental effects of frailty on alloHCT outcomes, the specific activity and sleep patterns associated with varying degrees of frailty—not frail, pre-frail, and frail—remain unexplored, particularly in myeloablative alloHCT settings among otherwise fit adults. Our pilot study aims to investigate these patterns, potentially guiding more personalized care to improve quality of life and survival for alloHCT recipients.

## METHODS

### Patient Selection

We included adults (≥18 years) scheduled for their first myeloablative alloHCT at the University of Minnesota who had a smartphone or tablet compatible with the Fitbit® app from June 2022 to January 2023. Participants received a Fitbit® Sense device before their alloHCT hospitalization and were advised to stay active by walking within the hospital unit. Access to physical therapists and exercise facilities was provided. The study adhered to IRB approval and HIPAA guidelines, with all participants giving written informed consent.

### Data Collection and Definitions

Participants synced their Fitbit® with a PHI-compliant Fitabase® at least weekly. We included data from admission until day +30 post-transplant, considering days with ≥10 hours of wear time as evaluable days. Activity was measured via daily steps, active and sedentary time—active time being periods with ≥3 METS energy expenditure. Sleep analysis included duration, deep sleep, and REM sleep. Frailty status, assessed pre-transplant using Fried Phenotype criteria (5), categorized individuals as frail (≥3 criteria), pre-frail (1-2 criteria), or not frail (no criteria), with data extracted from health records.

### Statistical Methods

Descriptive statistics illustrated Fitbit® data by frailty status. Differences between not frail, pre-frail, and frail groups were analyzed using Kruskal-Wallis tests and Dunn’s multiple comparisons for post hoc assessments. Smoothing spline regression curves show longitudinal activity and sleep trends. Statistical analyses were completed using JMP Pro 17 (SAS Corporation, Cary, NC) and GraphPad Prism version 9.4.1 (GraphPad Software, Boston, MA).

## RESULTS

### Patient Characteristics

Nine patients with adequate data were included in the analysis. Median age was 48 (range 22 to 59); five patients were male and four were female. Two patients were categorized as frail, five as pre-frail, and two as not frail prior to alloHCT. Additional patient characteristics are included in Supplemental Table 1. The mean percentage of evaluable days between hospital admission to day +30 was 81% ± 22% with a mean wear time of 1307 ± 198 minutes.

### Patterns of Activity

Not frail patients had a higher mean of daily steps compared to pre-frail (2.4-fold higher, 8265 ± 5274 versus 3381 ± 2757 steps, p < 0.0001) and frail (4.2-fold higher, 1977 ± 1610 steps, p < 0.0001). Pre-frail patients had a higher mean of daily steps than frail (1.7-fold higher, p = 0.0103, Figure 1a). Not frail patients also had a higher mean of active minutes compared to pre-frail (56 ± 49 versus 20 ± 26 minutes, p < 0.0001) and frail (25 ± 47 minutes, p < 0.0001). There was no significant difference in the mean of active minutes between pre-frail and frail patients (p>0.9999). Not frail patients had a lower mean of sedentary minutes compared to both pre-frail (763 ± 206 versus 884 ± 313 minutes, p = 0.0064) and frail (943 ± 422 minutes, p = 0.0004). There was no statistically significant difference in the mean sedentary time between pre-frail and frail patients (p=0.2472). Maximum sedentary time occurred around day +15 post-transplant (Figure 1c).

**Figure 1:**
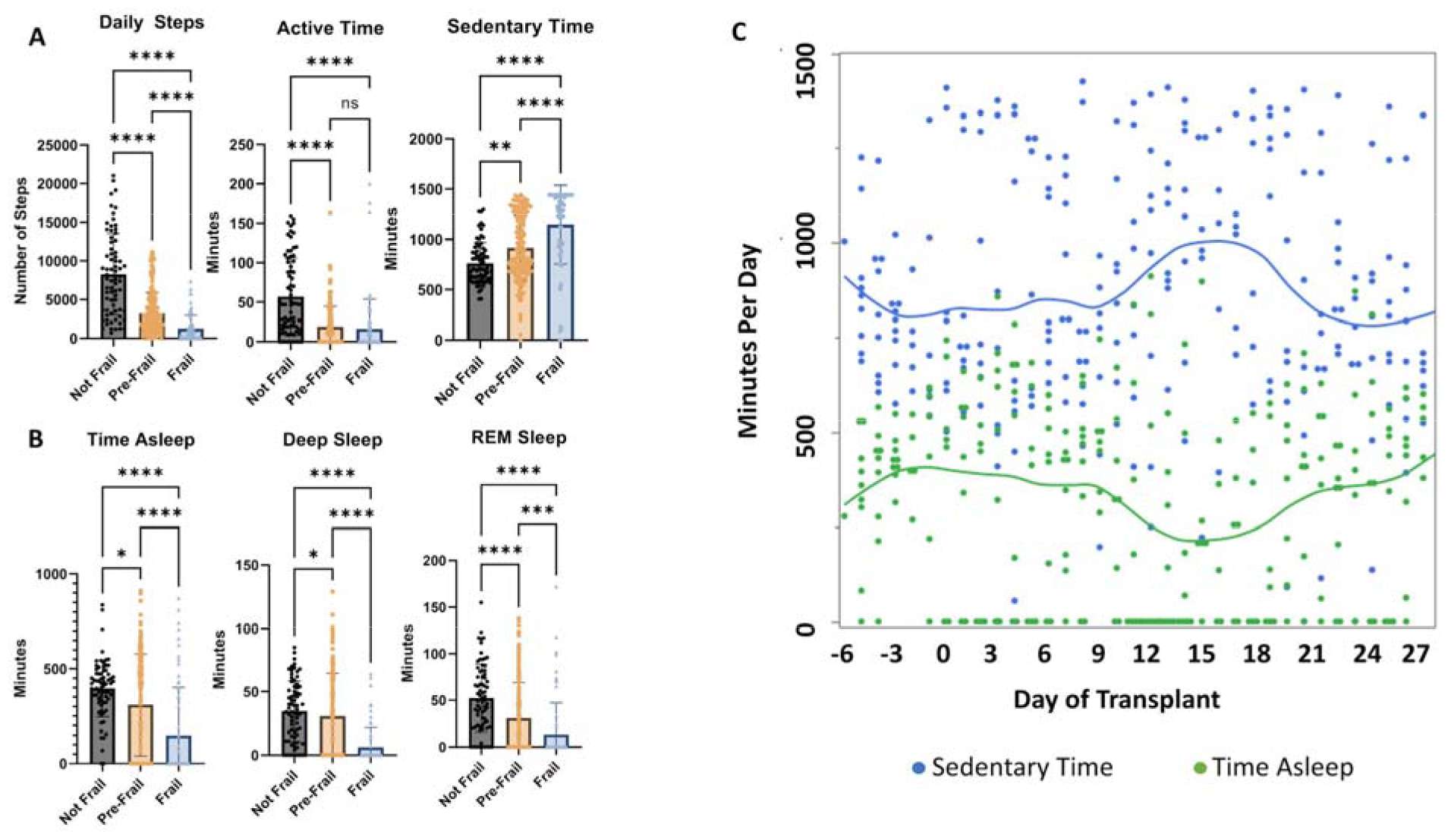
Patterns of (A) activity and (B) sleep based on phenotype of Frailty. The top of the bar represents the mean with error bars representing standard deviation. Longitudinal sedentary time and minutes asleep by transplant date are shown in C, with the line representing the smoothing spline regression curves.

### Patterns of Sleep

Across all cohorts, there was a median percentage of 67% (range 0% to 100%) of evaluable days including complete sleep data with sleep stages from admission to day +30. Overall, not frail patients had the best sleep quantity and quality (Figure 1b). Not frail patients spent more time in REM sleep compared to pre-frail (52 ± 36 versus 33 ± 39 minutes, p = 0.0003) and frail (24 ± 43 minutes, p < 0.0001). Frail patients spent less time in deep sleep compared to pre-frail (11 ± 19 versus 33 ± 34 minutes, p = 0.0003) and not frail (35 ± 24 minutes, p < 0.0001). There was a median percentage of 28% (range 0% to 58%) of days with zero minutes of time asleep, a median percentage of 36% (range 3% to 87%) of days with zero minutes of time in deep sleep, and a median percentage of 36% (range 0% to 87%) of days with zero minutes of REM sleep as measured by the Fitbit®. The sleep time nadir was also around day +15 post-transplant (Figure 1c).

## DISCUSSION

Our study delineates the variances in activity and sleep patterns among three distinct frailty phenotypes prior to transplantation. We discovered that patients without frailty exhibit higher daily step counts and more active time than their pre-frail and frail counterparts. Furthermore, individuals classified as frail not only engaged in less physical activity but more surprisingly also experienced diminished durations of restorative deep and REM sleep. The occurrence of days without recorded sleep data suggests that sleep disruptions are prevalent across all patient groups. Such disturbances may be partly explained by the Fitbit®’s methodology, which only logs sleep after an hour without movement and requires a three-hour minimum of sleep data to analyze sleep stages (6, 7). The frequent nighttime awakenings, as reported by El Jurdi et al., averaging 4.5 interruptions per night, are a probable factor in the poor sleep quality captured in our findings (4).

Previous research has linked frailty to decreased overall survival, increased non-relapse mortality, and a decline in the quality of life (2, 3, 8, 9). The reduced activity and lack of restorative sleep observed in frail patients are likely contributing to these negative health outcomes. Interventions tailored to the specific aspects of frailty in the elderly have been shown to enhance physical condition, as noted by Cameron et al. (10). This insight opens the possibility of applying similar targeted interventions to alloHCT patients, employing pre-transplant frailty assessments to inform and customize therapeutic strategies, potentially mitigating the impact of frailty on post-transplant recovery and long-term well-being.

## CONCLUSION

Our study demonstrated a reduced level of physical activity and poor sleep in individuals with frailty undergoing myeloablative alloHCT. Recognizing these differences may allow for development of targeted interventions to reduce disease burdens of care and improve support systems.

## Data Availability

Reasonable requests for data can be sent to the corresponding author.

## Notes

### Competing Interest Statement

The authors have declared no competing interest.

### Funding Statement

This study was funded by the Division of Hematology, Oncology, and Transplantation, Department of Medicine, University of Minnesota.

### Author Declarations

This study (STUDY00011670) was approved by the University of Minnesota Institutional Board Review.

