## Supplemental Table 1 for "Frailty is associated with low physical activity and poor sleep quality in patients undergoing myeloablative allogeneic hematopoietic cell transplantation: A Fitbit® Pilot Study"

| Participant | Diagnosis | Age category  (years) | Sex | Donor type | Frailty status with criteria met | Fat/Lean Mass Ratio | Bone Density | Length of hospitalization (days) | Complications during hospitalization | Day 100 relapse and mortality outcomes |
| --- | --- | --- | --- | --- | --- | --- | --- | --- | --- | --- |
| 1 | Mantle cell lymphoma | 40-50y | M | Matched related donor | Not frail | 0.249 | 1.335 | 24 | ▪Mucositis ▪Malnutrition requiring total parenteral nutrition | Relapse free |
| 2 | Acute myeloid leukemia | 20-30y | M | Matched related donor | Frail ▪Weight loss ▪Exhaustion ▪Weakness | 0.380 | 1.123 | 27 | ▪Mucositis ▪Malnutrition requiring total parenteral nutrition | Relapse-related mortality |
| 3 | Acute lymphoblastic leukemia | 50-60y | F | Matched unrelated donor | Pre-frail ▪Weight loss ▪Weakness | 0.697 | 1.098 | 36 | ▪Mucositis ▪Malnutrition requiring total parenteral nutrition | Relapse free |
| 4 | Chronic myeloid leukemia | 40-50y | M | Matched unrelated donor | Not frail | 0.268 | 1.169 | 25 | ▪Mucositis ▪Neutropenic fever | Relapse free |
| 5 | Acute myeloid leukemia | 50-60y | F | Matched unrelated donor | Pre-frail ▪Exhaustion ▪Weakness | 0.468 | 0.956 | 25 | ▪Mucositis ▪Malnutrition requiring total parenteral nutrition ▪Clostridium difficile | Relapse free |
| 6 | Acute lymphoblastic leukemia | 20-30y | M | Matched related donor | Pre-frail ▪Weakness | 0.474 | 1.096 | 33 | ▪Mucositis ▪Neutropenic fever ▪Non-neutropenic fever of unknown source ▪Anterior mediastinal hematoma ▪Malnutrition requiring total parenteral nutrition | Relapse free |
| 7 | Myelodysplastic syndrome | 50-60y | F | Matched unrelated donor | Pre-frail ▪Weakness ▪Slow walking speed | 0.564 | 1.042 | 27 | ▪Mucositis ▪Neutropenic fever | Relapse, alive |
| 8 | Acute lymphoblastic leukemia | 30-40y | M | Matched related donor | Frail ▪Weight loss ▪Weakness ▪Low physical activity | 0.502 | 1.113 | 24 | ▪Mucositis ▪Neutropenic fever ▪Bacteremia | Relapse free |
| 9 | Acute myeloid leukemia | 50-60y | F | Matched unrelated donor | Pre-frail ▪Exhaustion ▪Weakness | 0.539 | 1.026 | 27 | ▪Mucositis ▪Neutropenic fever ▪Malnutrition requiring total parenteral nutrition | Relapse free |
